# Construction and empirical of ICU patient follow-up model based on symptom management theory: a quasi-randomized controlled trial study protocol

**DOI:** 10.1101/2024.04.03.24305306

**Authors:** Qinqin Li, Li Yao, Tingshu Wang, Tingrui Wang, Yan Liu

## Abstract

**Introduction:** With the gradual improvement of medical treatment and nursing, more and more patients are successfully transferred out of the ICU. However, intensive care unit (ICU) survivors often experience long-term physical, cognitive, and psychological problems, and their family members also experience physical and psychological dysfunction, summarized as post-intensive care syndrome (PICS), affecting their health-related quality of life. Post-ICU follow-up can improve post-ICU syndrome in patients and their families, but the optimal mode of post-ICU follow-up remains uncertain. The purpose of this study was to build a follow-up model of ICU patients based on symptom management theory.

**Methods and analysis:** This study is a quasi-randomized controlled trial that will be conducted in multiple ICUs at one hospital in China, with patients enrolled from January 2024 to December 2024 and followed for 6 months. We expect to enroll 180 ICU patients. The primary outcome measure was the incidence of post-ICU syndrome (PICS) after ICU transfer, and the secondary outcome measure was the incidence of PCS-F, health economics evaluation, and patient/family satisfaction.

**Ethics and dissemination:** The protocol was approved by the research ethics committee of the Affiliated Hospital of Guizhou Medical University (2023 ethical review No. 061). The results will be published in peer-reviewed journals and presented at national and international scientific conferences to publicize the research to healthcare professionals, health service authorities and the public.

**Strengths and limitations of this study:**

1. The strength of this study is that this project adopts systematic and scientific research methods, and the ICU post-follow-up model built on the basis of symptom management theory aims to provide patients with the required, predictable and whole-course ICU post-follow-up service, which is helpful to meet the needs of patients under the realistic situation of limited medical resources, and is conducive to improving the utilization efficiency of medical resources. It is of great significance to optimize medical service system.
2. In the process of clinical implementation of randomized controlled trials, there are many resistance and force majeure factors, and ICU patients are especially special, and they are still in a weak state after being transferred out of ICU, and the implementation of randomized controlled trials is very difficult. Based on this, this study adopts experimental research.
3. This study was only conducted in one hospital in Guizhou, China, which may have some limitations and should be expanded in the future.
4. The nature of the intervention does not allow blinding of study personnel and eligible patients at ICUs.

## Introduction

In recent years, there has been a notable worldwide surge in the population of individuals who have successfully emerged from intensive care units (ICUs), owing to the progress made in the fields of intensive care and nursing ^[1]^. Multiple studies have provided evidence that individuals who have undergone critical illness frequently encounter the development or worsening of various functional impairments, including physical, cognitive, psychological, and social consequences, upon their discharge from the intensive care unit (ICU)^[2,3]^. Simultaneously, the admission of a patient to the ICU and the subsequent caregiving responsibilities impose a range of pressures on the family, resulting in physical and psychological dysfunction among its members, commonly known as the Post-intensive Care Syndrome Family (PICS-F)^[4]^.These phenomena are collectively recognized as Post-intensive Care Syndrome (PICS) ^[5,6]^. Following the manifestation of Post-Intensive Care Syndrome (PICS), there arises an increased need for medical resources, an augmented societal burden, an extended period of patient recuperation, hindered reintegration into society, academic pursuits, and employment, as well as a diminished overall quality of life^[7,8]^. Some researchers have even identified post-ICU syndrome (PICS) as a public health problem that needs to be addressed^[9]^. The United Kingdom of Great Britain and Northern Ireland(UK) National institute for Health and Care Excellence (NICE) guidelines recommend that for patients with or with potential risk factors for PICS, it is essential to develop a comprehensive long-term complete rehabilitation plan ^[10]^. The conference on survivors of intensive care units (ICUs) organized by the Society of Critical Care Medicine (SCCM) highlighted the importance of not only increasing awareness of Post-Intensive Care Syndrome (PICS) among both the general public and medical professionals, but also investigating specific preventive and intervention strategies for PICS. One intervention that received considerable attention was the implementation of post-ICU follow-up services^[11]^.

Post-intensive care unit (ICU) interventions encompass the implementation of ongoing care protocols during the transition from the ICU and subsequent discharge, which are commonly known as post-ICU follow-up services^[12]^. Post-ICU follow-up has been shown to improve the outcomes of patients discharged from the ICU, but the overall quality remains to be improved^[13,14]^. In a longitudinal study spanning three years, it was observed that the implementation of a multidisciplinary collaborative model of intensive care unit (ICU) post-specialty clinic significantly mitigated the occurrence of sequelae, including psychological and cognitive impairments, among patients^[15]^. Furthermore, empirical researches ^[16,17]^ have demonstrated that post-ICU clinics led by nurses do not yield a substantial enhancement in the quality of life for individuals who have survived an intensive care unit stay, even after one year of discharge. Additionally, the outcomes of economic evaluations do not indicate a reduction in healthcare expenses for patients. Indeed, a universally applicable rehabilitation intervention that can enhance the functional capacity of individuals who have survived an intensive care unit (ICU) stay does not exist^[18]^. Furthermore, the current post-ICU follow-up services lack a standardized framework. These services encompass: (1) Implementing a continuum of functional exercise-based interventions from the ICU to the general ward and extending beyond discharge, which includes cognitive training, supervised intensive training, information provision, and psychological intervention^[19]^. (2) Establishing an ICU post-clinic that offers a comprehensive rehabilitation program tailored to the needs of patients^[20]^. (3) Implementing an ICU diary is recommended to enhance the exchange of information and promote the psychological recovery of patients following their discharge from the intensive care unit (ICU) ^[21]^.

To enhance the long-term prognosis of critically ill individuals, the establishment of a follow-up model for patients in the intensive care unit holds considerable importance^[22]^. This research endeavor aims to construct a follow-up model for ICU patients, based on the principles of symptom management theory. Symptom Management Theory (SMT) is a theory situated within the mid-domain and characterized by its process-oriented nature, originating from the Symptom Management Model (SMM)^[23]^. SMT elucidates the multifaceted components and interconnectedness entailed in a proficient symptom management procedure^[24]^. SMT identified seven outcome measures associated with symptom change: functional status, emotional status, self-care, medical costs, quality of life, morbidity rates, complications, and mortality rates^[25]^. Miaskowski^[26,27]^ suggested that in order to alleviate the burden associated with assessing multiple outcome indicators on patients and their families, it is advisable to restrict the number of indicators and instead focus on selecting one or a few indicators that are closely aligned with the study at hand. Based on an extensive review of relevant literature, this study identified three indicators that are highly pertinent to Post-Intensive Care Syndrome (PICS) and effectively capture the overall condition of patients: cognitive level, psychological status, and physiological function status. The selection of psychological and physiological states, deemed most pertinent to PCS-F, was made in order to accurately depict the condition of family members of patients in the intensive care unit. The theoretical framework pertaining to symptom management is ultimately established in this study, as depicted in Figure 1.

**Figure 1.**
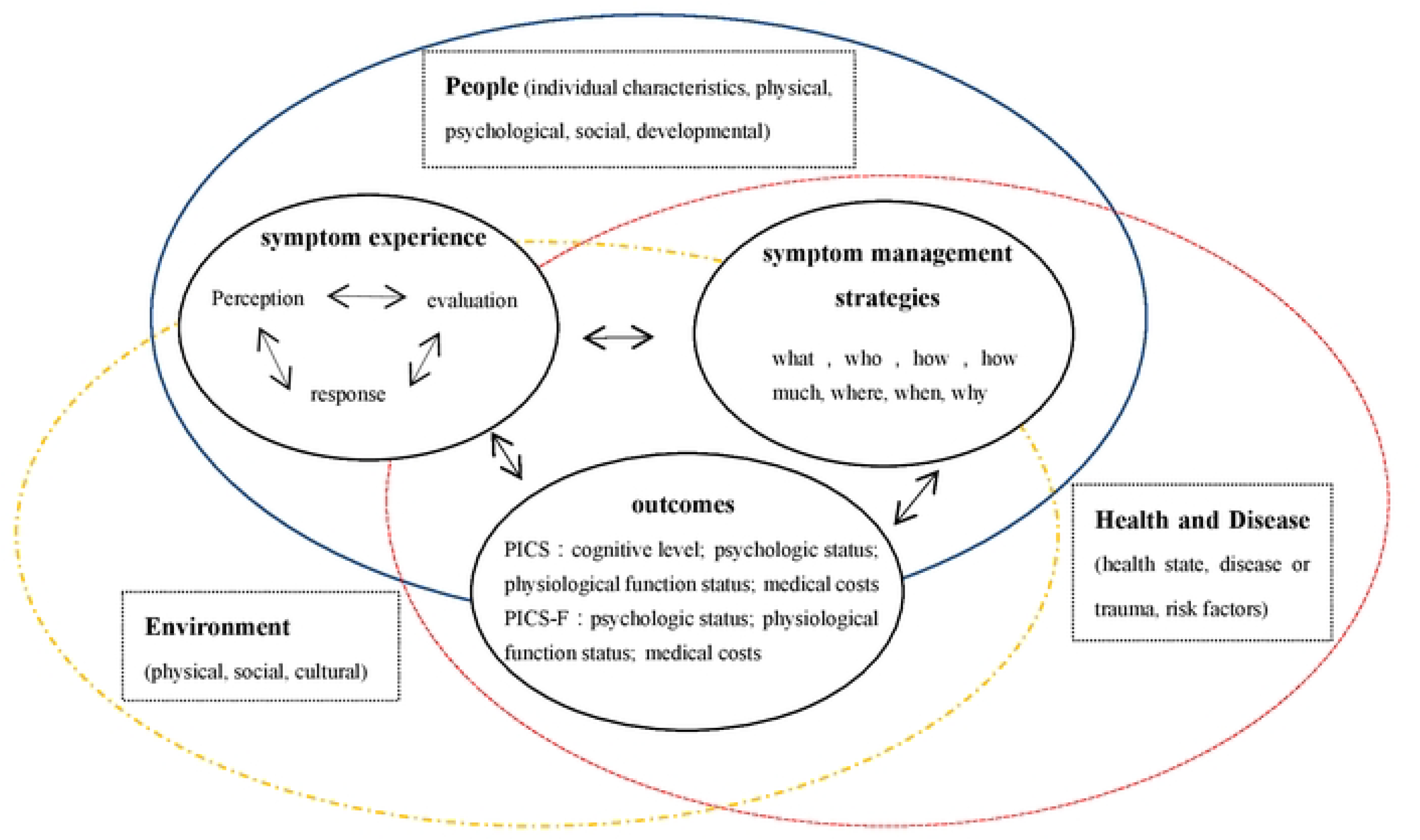
PICS and PICS-F symptom management models.

## Methods and analysis

### Aim and Objective

The objective of this study was to develop a post-ICU follow-up model grounded in symptom management theory in order to address the requirements of patients and their families. Additionally, the study seeks to investigate the effectiveness of this post-ICU follow-up model through a quasi-randomized controlled trial, specifically focusing on enhancing the functional, psychological, and cognitive well-being of critically ill patients following their discharge from the ICU. It is hypothesized that the intervention group will experience a significantly greater reduction in the occurrence of post-ICU syndrome compared to the control group.

### Study design and setting

This study will employ a quasi-randomized controlled trial to explore the effectiveness of post-ICU follow-up model based on symptom management theory. The research will be carried out within the ICU of a hospital located in Guizhou, an inland province in China. The study site encompasses an integrated ICU, Respiratory Intensive Care Unit (RICU), Cardiac Care Unit (CCU), and Medical Intensive Care Unit (MICU), boasting a collective capacity of 84 beds and an annual transfer of approximately 900 patients from the ICU. The hospital’s integration of teaching and scientific research, along with its collaborative relationships with several secondary hospitals and communities, ensures a reliable foundation for the successful execution of this study. ICU patients will be recruited between January 2024 and December 2024 with a subsequent follow-up for 6 months. To mitigate the impact of contamination and seasonal variables, we will employ the time grouping technique, wherein the experimental group will consist of the sample size collected during the first half of each month, while the control group will comprise the sample size collected during the second half of each month. This study has obtained approval from the Ethics Committee of the hospital, bearing the approval number 2021 Ethics Review No. 064.

### Study population and eligibility criteria

We aim to include 180 ICU patients, and patients admitted to the ICU at the participating site will be routinely screened against the following eligibility criteria. Patients are eligible to participate when they are 18 years or older; admitted at least 24 hours to ICU; ICU treatment results in survivors and gave written informed consent (or by their legal representative). Patients are not eligible for the study when they are less than 18 years or older than 80 years; with severe vision and hearing impairment (who cannot complete the scale test or undergo follow-up); with significant disability or dysfunction, such as amputation or fistula; receive palliative care; dropped out of the trial.

### Patient recruitment

Prior to trial commencement, participating sites have to provide informed consent on an institutional level. Patients will be identified and screened for eligibility by the local team in the ICU. Patients will receive information from ICU nurses and intensivists regarding the aim, content and relevance of the study, and will be asked for participation. Informed consent is asked for the questionnaires and data from the patients’ individual hospital information system (HIS). Written informed consent is obtained by the patient or the legal representative if the patient is unable to consent.

### Sample size calculation

The formula for conducting two independent sample rate comparisons was employed to determine the incidence of Post-Intensive Care Syndrome (PICS) as the calculation index.

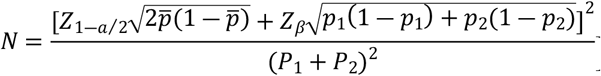 In the given formula, N represents the sample size required for each group, *p*1 denotes the incidence rate of the control group, and *p*2 signifies the incidence rate of the experimental group, 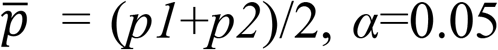, *α*=0.05, *β*=1%, and the certainty (*1-β*) is 99%, referring to relevant literature, *p*_1_=18.61%, *p*_2_=4.65%^[28]^. It is concluded that the number of required samples in each group is 82, consider a 10% exit rate, the number of required samples in each group is 90, and the total sample size is 180.

### Intervention

#### Usual Care

The study will compare the intervention with standard care. Upon admission, patients will be transferred to the intensive care unit (ICU) following a definitive diagnosis and will receive appropriate symptomatic treatment and nursing interventions. Once the patient’s condition becomes stable and meets the criteria for ICU discharge, they will be transferred to a general ward. The control group will receive routine nursing interventions, such as monitoring vital signs, providing dietary and exercise guidance, offering medication instructions, delivering health education, and providing discharge instructions. The experimental group will be based on the control group. On the basis of the control group, the experimental group was given the pre-constructed ICU follow-up mode intervention.

### ICU Patient Follow-up Model

Patients who meet the inclusion criteria are transferred from the ICU to the general ward and will receive the usual care and ICU patient follow-up intervention, and their families will also receive the ICU patient follow-up intervention. The detailed content of the post-ICU multidisciplinary team follow-up model is given in Table 1. This follow-up model was formulated by a multidisciplinary team comprising five nursing specialists, two senior intensivists, one psychiatrist, and two chronic disease management experts. Each member of this multidisciplinary team possesses a minimum of ten years of clinical nursing experience and substantial expertise in clinical management. The construction process of ICU Patient Follow-up Model is shown in Figure 2. Before constructing the follow-up model, we will perform the following tasks: firstly, we will develop the Post-intensive Care Syndrome Family Questionnaire (PICSQ-F). Based on the tool and the post-intensive Care Syndrome Questionnaire (PICSQ) developed by Jeong, a Korean scholar, translated by Tingting Sun^[29]^, We will conduct a cohort studies to explore the occurrence and influencing factors of PICS and PICS-F on the day of discharge and at 1, 3, and 6 months after discharge. Secondly, we will use the self-prepared “Family/Patient Post-ICU Health Needs Questionnaire” to investigate the health needs of ICU patients and their families, and use semi-structured interview method to further explore the deep-level needs of family members and ICU patients.

**Table 1.**
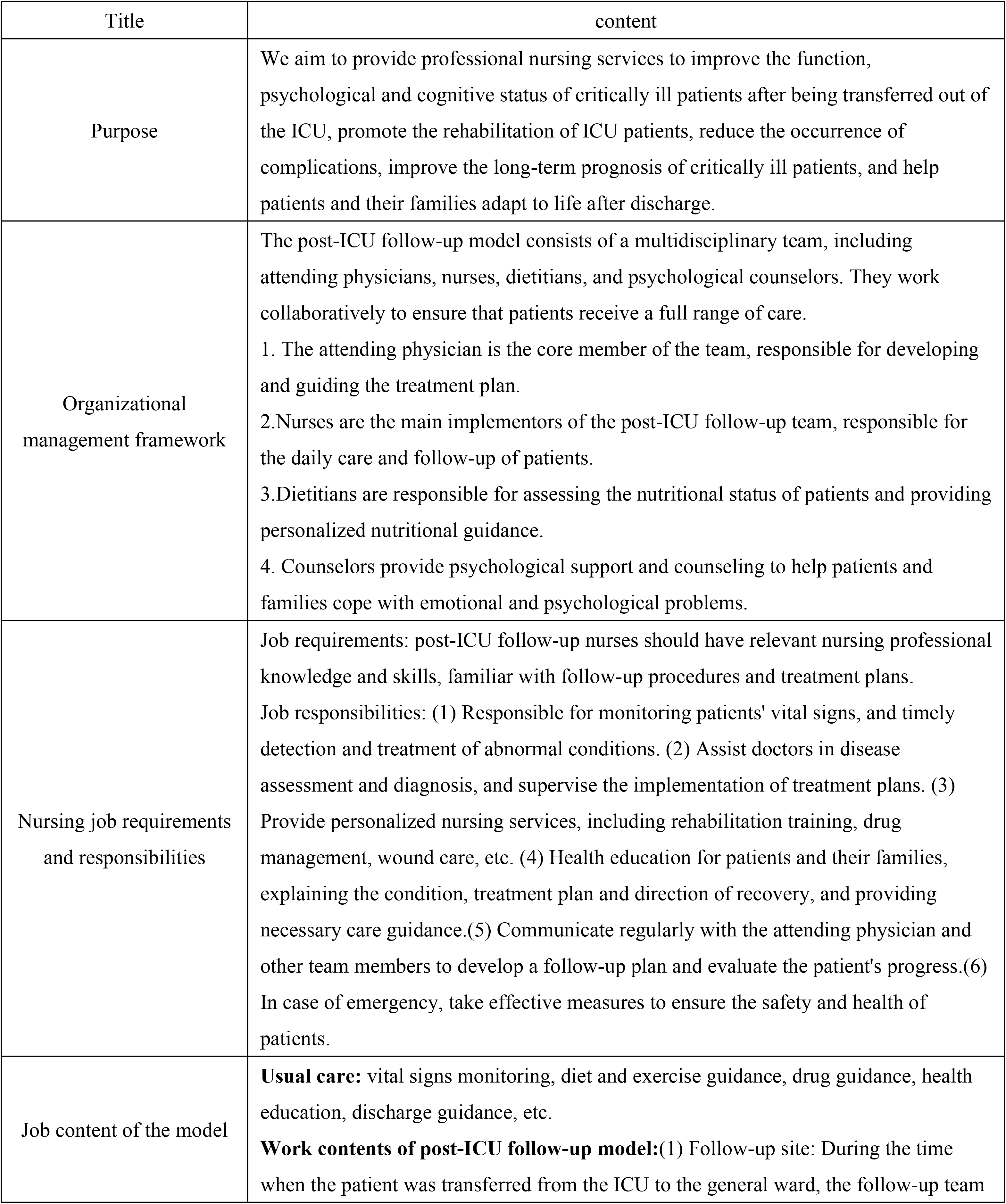
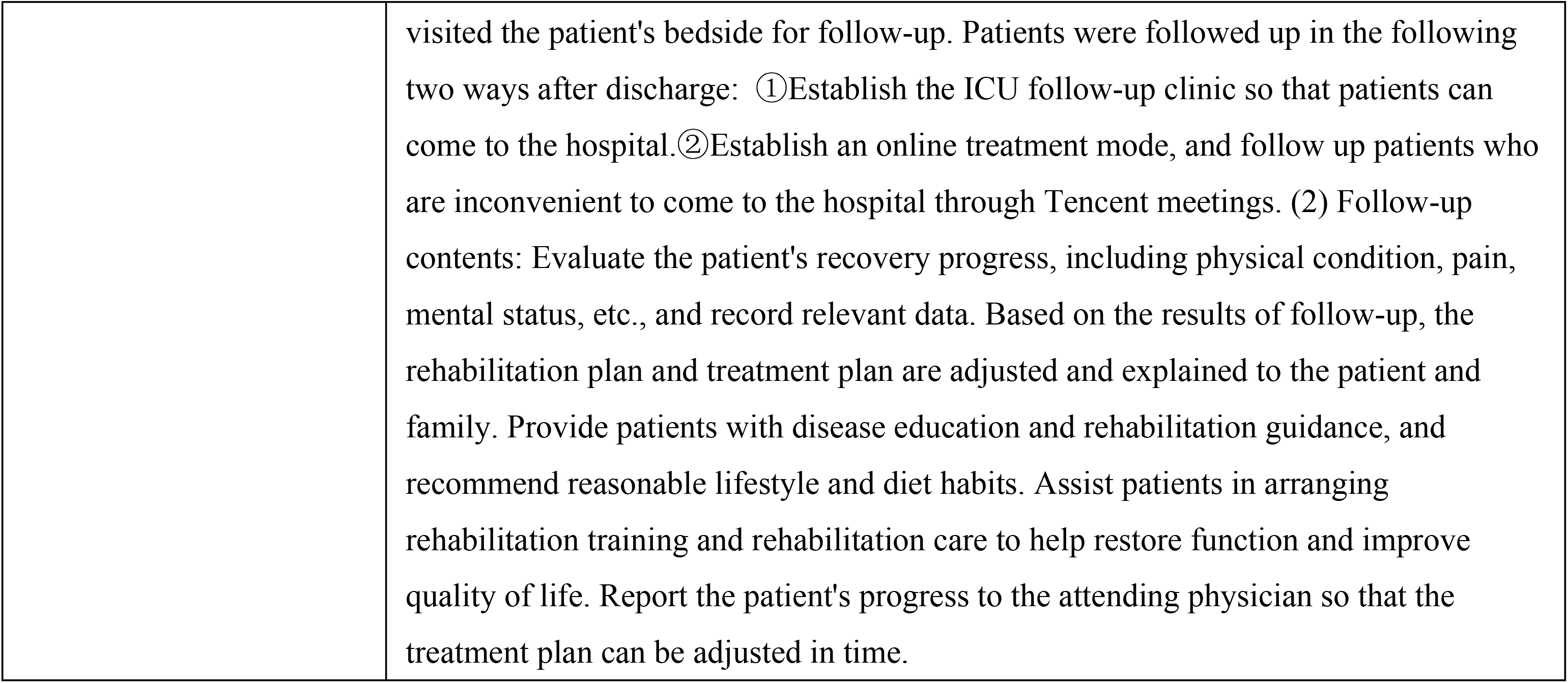
Detail content of ICU patient follow-up model

| Title | content |
| --- | --- |
| Purpose | We aim to provide professional nursing services to improve the function, psychological and cognitive status of critically ill patients after being transferred out of the ICU, promote the rehabilitation of ICU patients, reduce the occurrence of complications, improve the long-term prognosis of critically ill patients, and help patients and their families adapt to life after discharge. |
| Organizational management framework | The post-ICU follow-up model consists of a multidisciplinary team, including attending physicians, nurses, dietitians, and psychological counselors. They work collaboratively to ensure that patients receive a full range of care.<br>1. The attending physician is the core member of the team, responsible for developing and guiding the treatment plan.<br>2. Nurses are the main implementors of the post-ICU follow-up team, responsible for the daily care and follow-up of patients.<br>3. Dietitians are responsible for assessing the nutritional status of patients and providing personalized nutritional guidance.<br>4. Counselors provide psychological support and counseling to help patients and families cope with emotional and psychological problems. |
| Nursing job requirements and responsibilities | Job requirements: post-ICU follow-up nurses should have relevant nursing professional knowledge and skills, familiar with follow-up procedures and treatment plans.<br>Job responsibilities: (1) Responsible for monitoring patients' vital signs, and timely detection and treatment of abnormal conditions. (2) Assist doctors in disease assessment and diagnosis, and supervise the implementation of treatment plans. (3) Provide personalized nursing services, including rehabilitation training, drug management, wound care, etc. (4) Health education for patients and their families, explaining the condition, treatment plan and direction of recovery, and providing necessary care guidance. (5) Communicate regularly with the attending physician and other team members to develop a follow-up plan and evaluate the patient's progress. (6) In case of emergency, take effective measures to ensure the safety and health of patients. |
| Job content of the model | **Usual care:** vital signs monitoring, diet and exercise guidance, drug guidance, health education, discharge guidance, etc.<br>**Work contents of post-ICU follow-up model:** (1) Follow-up site: During the time when the patient was transferred from the ICU to the general ward, the follow-up team |
|  | visited the patient's bedside for follow-up. Patients were followed up in the following two ways after discharge: ①Establish the ICU follow-up clinic so that patients can come to the hospital.②Establish an online treatment mode, and follow up patients who are inconvenient to come to the hospital through Tencent meetings. (2) Follow-up contents: Evaluate the patient's recovery progress, including physical condition, pain, mental status, etc., and record relevant data. Based on the results of follow-up, the rehabilitation plan and treatment plan are adjusted and explained to the patient and family. Provide patients with disease education and rehabilitation guidance, and recommend reasonable lifestyle and diet habits. Assist patients in arranging rehabilitation training and rehabilitation care to help restore function and improve quality of life. Report the patient's progress to the attending physician so that the treatment plan can be adjusted in time. |

**Figure 2.**
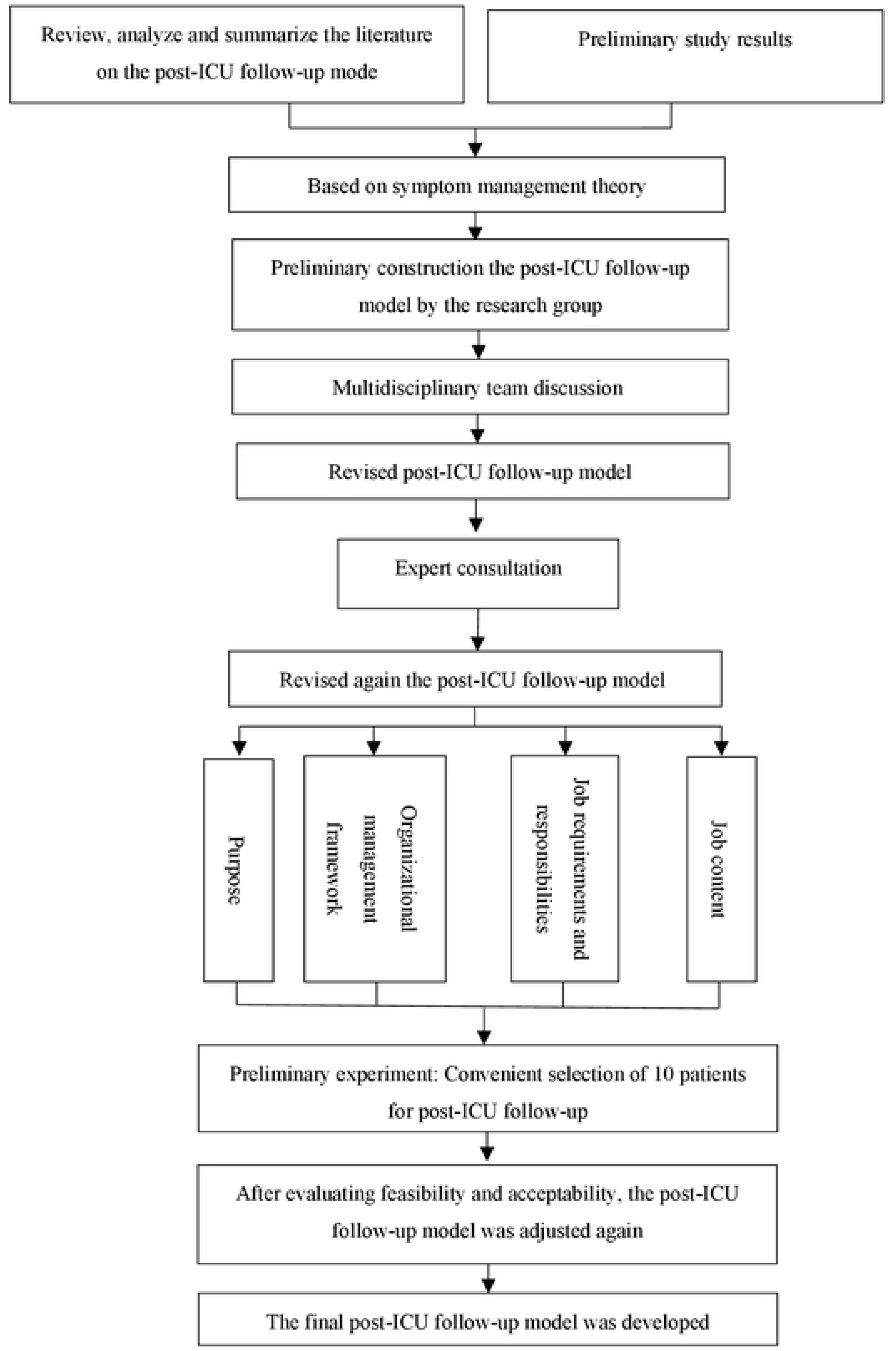
Flow diagram of the construction of ICU patient follow-up model.

### Study procedures

We will strictly screen the subjects according to inclusion and exclusion criteria, and the subjects will be divided into two groups, namely the intervention group and the control group, after which the baseline data of the patients will be collected. The control group will be given routine care for intervention, and the experimental group will be given routine care and pre-constructed ICU follow-up mode for intervention. Our follow-up period is 6 months. We conducted a midpoint assessment at 1 month of intervention implementation; Endpoints will be assessed at 6 months after the intervention. The assessment included the incidence of post-ICU syndrome (PICS), the incidence of family post-ICU syndrome (PICS-F), health economics evaluation, and patient/family satisfaction. The flow chart of the study procedures is shown in Figure 3.

**Figure 3.**
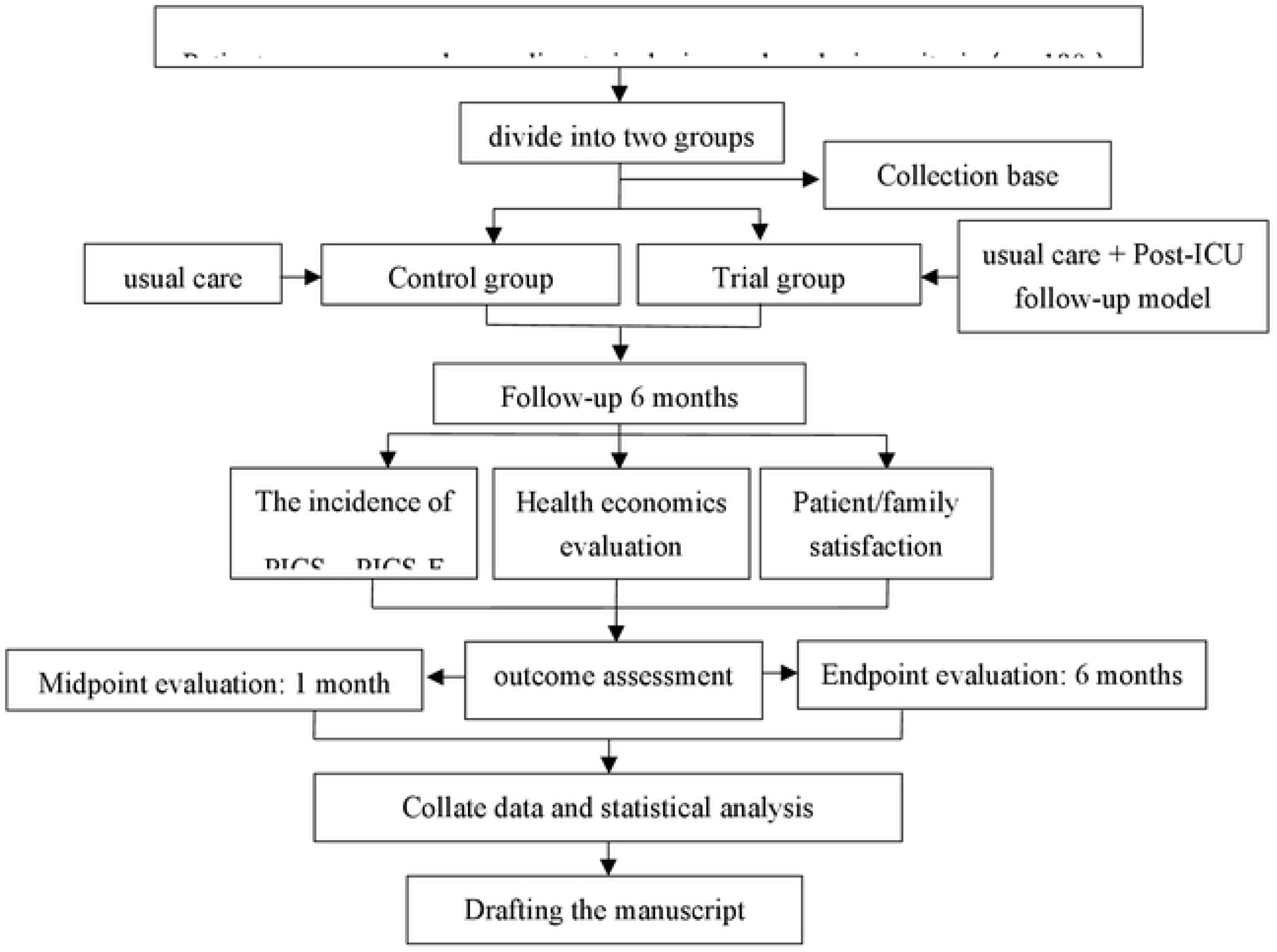
Flow chart of applied study of post-ICU follow-up model.

### Outcomes measures

The primary outcome measure is the incidence of post-ICU syndrome (PICS) in patients transferred from the ICU. The secondary outcome measures are patients’ the incidence of PICS-F, health economics evaluation, and patient/family satisfaction.

### Data collection

Different methods will be used to collect data among ICU patients. Before data collection, researchers were given uniform training on data collection methods. General data were collected using a self-designed general data questionnaire, including patient/family socio-demographic data and patient clinical disease treatment data; The collection time was after the patients were enrolled and before the intervention program was implemented; Data were extracted from the hospital’s HIS system and patients/family members were questioned. The occurrence of PICS was assessed using the Chinese-language PICSQ questionnaire by Sun Tingting et al^[29]^. The first assessment time was after the patients were enrolled and before the intervention program was implemented; The midpoint evaluation time was 1 month after the intervention; The end point was evaluated 6 months after the intervention. The health economics evaluation included hospitalization costs after ICU transfer, length of stay after ICU transfer, medical costs after discharge, degree of impact on post-discharge income (current income/previous income), European five-dimensional Health Scale, quality-adjusted life years, disability-adjusted life years. Family Satisfaction in the Intensive Care Unit(FS-ICU) was used to evaluate patient/family satisfaction, there were 24 items in the questionnaire, Cronbach’s α=0.90; All items were scored using likert 5-level score, and each item was converted into a value ranging from 0 to 100, the greater the value, the higher the satisfaction^[30]^.

### Statistical analysis

In this study, Excel 2023 will be used to establish a database and SPSS 25.0 statistical software will be used for statistical analysis of the data. In this study, the data will be recorded by two people and a random sample of 10% of the data will be cross-checked. Consult professional statistical analysts when analyzing data. Baseline demographics characteristics will be quantified using descriptive statistics. Continuous variables will be presented as mean (SD) or as median (95% range), based on the distribution of the variable. Categorical variables will be presented as absolute number and relative frequency.

### Patient and public involvement

Patients or the public were not involved in the design, or conduct, or reporting, or dissemination plans of our research.

### Ethics and dissemination

The study has been approved by the research ethics committee of the Medical Science Ethics Committee of the Affiliated Hospital of Guizhou Medical University, and the approval number is: 2023 ethical review No. 061. The principle of this study is voluntary. Patients or their family members will be informed of the purpose, significance and method of the study, and their consent and informed consent will be obtained. Signed informed consent will be obtained from all participants. On completion of the study, its findings will be published in peer-reviewed journals and presented at national and international scientific conferences to publicise the research to healthcare professionals, health service authorities and the public. A summary of the results will be made available to the study patients if requested.

## Data Availability

Deidentified research data will be made publicly available when the study is completed and published.

